# Worsening of kidney function is the major mechanism of heart failure in hypertension: the ALLHAT study

**DOI:** 10.1101/2020.06.15.20132241

**Authors:** Maedeh Khayyat-Kholghi, Suzanne Oparil, Barry R. Davis, Larisa G. Tereshchenko

## Abstract

**Background:** We aimed to quantify the extent to which the effect of antihypertensive drugs on incident heart failure (HF) is mediated by their effect on kidney function. We hypothesized that the dynamic change in kidney function is the mechanism behind differences in the rate of incident HF in ALLHAT participants randomized to lisinopril, amlodipine, and doxazosin, in comparison to those randomized to chlorthalidone.

**Methods:** Causal mediation analysis of ALLHAT data (1994-2002) included participants with available baseline and 24-48 month estimated glomerular filtration rate (eGFR) (n=27,918; mean age 66±7.4; 32.4% black, 56.3% men). Change in eGFR was the mediator. Incident symptomatic HF was the primary outcome. Hospitalized/fatal HF was the secondary outcome. Linear regression (for mediator) and logistic regression (for outcome) analyses were adjusted for demographics, cardiovascular disease, and risk factors.

**Results:** There were 1,769 incident HF events, including 1,359 hospitalized/fatal HF events. In fully adjusted causal mediation analysis, the relative change in eGFR mediated 38% of the effect of amlodipine, 25.5% of doxazosin, and 6.3% of lisinopril on incident symptomatic HF, and 42% of the effect of amlodipine, 55.3% of doxazosin, and 12.7% of lisinopril on hospitalized/fatal HF. In lisinopril arm, eGFR changes had an opposite effect on symptomatic versus hospitalized/fatal HF outcomes. Reduction in eGFR by at least 40% explained > 50% of increased risk in hospitalized/fatal HF but 18-25% reduction of symptomatic HF risk.

**Conclusion:** On the risk difference scale, change in eGFR accounts for more than 50% of the mechanism by which antihypertensive medications affect HF.

**Clinical Trial Registration:** URL:www.clinicaltrials.gov Unique identifier: NCT00000542.

## Introduction

Kidney dysfunction is a well-known risk factor for heart failure (HF),^1^carrying the highest predictive value among the HF risk scores.^2^ In patients with end-stage kidney disease (ESKD), and HF with reduced left ventricular ejection fraction (LVEF), kidney transplantation resulted in normalization of LVEF.^3^ Although it is well-accepted that hypertension accelerates kidney injury, the role of hypertension in the initiation of kidney disease remains controversial.^4^ While commonly used antihypertensive medications such as angiotensin-converting enzyme inhibitors (ACEIs) and diuretics may cause an increase in serum creatinine, this effect may be viewed as a favorable prognostic indicator for HF^5^ if associated with a decrease in signs and symptoms of congestion.^6^ In patients with hypertension, it is unclear to what degree impaired kidney function is a marker of HF severity or a reflection of a mechanism contributing to HF progression.^7^ The diagnosis, pathophysiology, prognosis, and treatment of the cardiorenal syndrome^1^ are focused on established chronic kidney disease (CKD) and HF. In contrast, knowledge gaps persist in understanding of the role of impaired kidney function as a mechanism that predisposes to the development of incident HF in hypertensive patients.

To address these knowledge gaps, we analyzed data from the Antihypertensive and Lipid-Lowering Treatment to Prevent Heart Attack Trial (ALLHAT).^8^ ALLHAT was a multicenter, randomized, double-blind trial designed to compare cardiovascular (CV) outcomes in high-risk hypertensive patients assigned to the ACEI lisinopril, the calcium channel blocker (CCB) amlodipine, and the 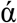-blocker doxazosin, in comparison to the thiazide-like diuretic chlorthalidone. We quantified the extent to which the effects of lisinopril, amlodipine, doxazosin, and chlorthalidone on incident HF are mediated by their effects on kidney function. We hypothesized that a dynamic change in kidney function is the mechanism behind previously observed differences in the rates of incident HF in ALLHAT participants randomized to lisinopril, amlodipine, and doxazosin, in comparison to those randomized to chlorthalidone.^9, 10^

## Methods

We used the ALLHAT dataset that is publicly available from the National Heart, Lung, and Blood Institute, via BioLINCC.^11^ The Oregon Health & Science University Institutional Review Board reviewed the study and determined the deidentified nature of the publicly available dataset.

### Study population

ALLHAT^8^ was conducted from 1994 to 2002. Adults age 55 and above with hypertension and at least one additional cardiovascular disease risk factor were enrolled. The risk factors included documented coronary heart disease (CHD), type 2 diabetes mellitus, left ventricular hypertrophy (LVH) on ECG or echocardiogram, smoking, high-density lipoprotein (HDL) < 35mg/dL, and ST-T ECG changes indicative of ischemia. Exclusion criteria included symptomatic HF or LVEF <35%, recent myocardial infarction (MI), stroke, poorly controlled hypertension, and a serum creatinine level >2 mg/dL (>176.8 μmol/L).^12^

In the current study, we included ALLHAT participants with available data on estimated glomerular filtration rate (eGFR) changes during in-trial follow-up. We excluded participants with missing eGFR follow-up data or missing covariates. The final study population (Figure 1) included 27,918 participants: 10,487 were randomized to chlorthalidone, 5,388 to doxazosin, 6,166 to amlodipine, and 5,877 to lisinopril.

**Figure 1.**
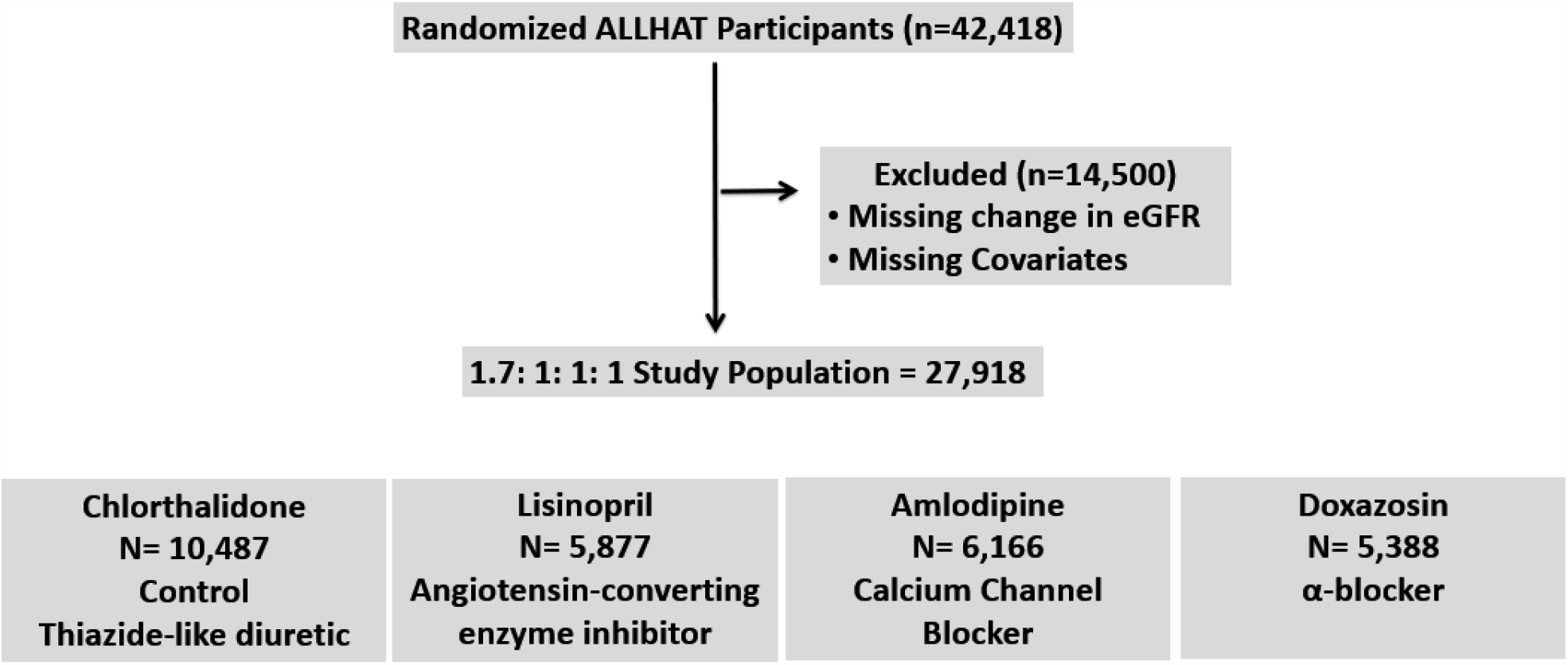
Study flow diagram.

### Mediator: eGFR change

As previously described^12^, at baseline, serum creatinine was measured at a central laboratory using the VITROS chemistry system (Ortho-Clinical Diagnostics, Rochester, NY) with a coefficient of variation of approximately 2%. Creatinine measurements were externally validated^13^ in two other laboratories, indicating that serum creatinine values measured in ALLHAT were comparable to the MDRD study measurements. Thus, use of the MDRD study equation in ALLHAT was justified.^12^

Estimated GFR was calculated using the simplified MDRD equation^12^: eGFR(mL/min/1.73m^2^)= 186.3 * (creatinine)^-1.154^ * (age)^-0.203^ * 1.212(Black) * 0.742(female).

Serum creatinine measurements were repeated at one month, one year, two years, and then every other year of follow-up.^14^ To calculate eGFR change during the trial, we subtracted baseline eGFR from the eGFR measured after at least 48 months (4 years) of follow-up. If 48-month eGFR measurement was not available, we subtracted baseline eGFR from the eGFR measured after at least 24 months (2 years) of follow-up. Participants with missing follow-up eGFR measurements were excluded. Next, we normalized eGFR change by the baseline eGFR value, to obtain relative eGFR change.

### Outcome: Incident heart failure

The primary outcome in the current study was incident symptomatic congestive HF, as defined by the ALLHAT investigators. Symptomatic congestive HF was diagnosed in the presence of both: (1) Paroxysmal nocturnal dyspnea, dyspnea at rest, New York Heart Association class III symptoms or orthopnea, and (2) rales, ankle edema (2+ or greater), sinus tachycardia of 120 beats per minute or more after 5 minutes at rest, cardiomegaly by chest X-ray, chest X-ray characteristic of congestive HF, S3 gallop or jugular venous distention. The ALLHAT HF validation study validated the incident HF outcome.^15^ In the current study, the secondary outcome was hospitalized/fatal HF. Both outcomes were validated by the ALLHAT HF validation study.^15-17^

### Covariates

The ALLHAT investigators obtained the baseline medical history by a combination of chart review and questioning during a routine office visit. The hypertension history included categories of participants who were treated for at least two months, or less than two months/untreated. Baseline CHD history included known MI (including silent MI), angina, cardiac arrest, angiographically defined coronary stenosis more than 50%, reversible perfusion defects on cardiac scintigraphy, or prior coronary revascularization procedures. History of revascularization included a history of angioplasty, stenting, atherectomy, bypass surgery (coronary; peripheral vascular; carotid; vertebrobasilar), or aortic aneurysm repair. The presence of major ST-segment depression or T wave elevation on an ECG in the past two years was recorded. History of other atherosclerotic CVD included documented peripheral arterial disease or cerebrovascular disease. Type II diabetes was defined as fasting plasma glucose > 140 mg/dl [7.77 mmol/L] or non-fasting plasma glucose >200 mg/dl [11.1 mmol/L] in the past two years and/or current treatment with insulin or oral hypoglycemic agents. History of HDL cholesterol < 35 mg/dl (0.91 mmol/l) on two or more determinations within the past five years was identified. The current smoking history was included. Baseline ECG-LVH was based on any ECG within the past two years, as previously described.^18^ Echocardiographic LVH (Echo-LVH) was defined as a combined wall (posterior wall plus interventricular septum) thickness ≥ 25 mm on any echocardiogram in the past two years.

Baseline blood pressure was assessed as an average of two blood pressure determinations taken at least one day apart, with each determination being an average of 2 measurements. At every visit (every three months for the 1^st^ year and every four months thereafter), blood pressure was recorded as an average of two measurements. As previously described^18^, we estimated the blood pressure-lowering by subtracting baseline blood pressure from the blood pressure obtained at the latest in-trial study visit available at year sixth, fifth, fourth, third, second, or first.

### Statistical analysis

Antihypertensive treatment groups were defined per intention-to-treat (ITT) randomization assignment. After confirmation of normal distribution, continuous variables are reported as means±standard deviation (SD) or as the median and interquartile range (IQR) if skewed distribution. For unadjusted comparison of clinical characteristics in participants with three tertiles of relative eGFR change, we used ANOVA and χ^2^test. To determine the association of clinical characteristics with relative eGFR change, we used multivariable linear regression models adjusted for age, sex, and race/ethnicity.

We conducted causal mediation analysis^19^, using counterfactual definitions of direct and indirect effects in parametric regression models, as implemented by VanderWeeleet al^20^, which allows for treatment-mediator interaction. We estimated two models: (1) a linear model for the mediator conditional on treatment and covariates, and (2) a logistic model for the outcome conditional on treatment, the mediator, and covariates. In this study, treatment randomization eliminated exposure-outcome and exposure-mediator confounding. Relative change in eGFR over the course of the trial was used as a mediator. We adjusted for mediator-outcome confounders^16, 21^: demographic (age, sex, race, and ethnicity) and clinical characteristics known to be associated both with kidney function and HF: levels of baseline systolic and diastolic blood pressure, in-trial systolic and diastolic blood pressure lowering as previously defined,^18^ length of antihypertensive treatment before enrollment, ECG- or echo-LVH, history of MI, stroke, or other CVD, coronary revascularization, major ST depression or T-wave inversion, HDL<35 mg/dL twice in the past five years, BMI, smoking, diabetes, use of aspirin, participation in the lipid-lowering ALLHAT trial, baseline levels of total cholesterol, eGFR, and potassium, and geographic region (East, Midwest, South, West, Canada, and Puerto Rico/Virgin Islands).

We calculated four types of effects: the total, natural direct, natural indirect, and controlled direct effects. Natural direct effect represents the influence of antihypertensive treatment that is independent of eGFR changes, in the absence of eGFR changes (e.g., via blood pressure-lowering or drug-specific pharmacodynamics). A natural indirect (mediated) effect represents the influence of an antihypertensive drug that can be explained by its influence on dynamic eGFR changes over the course of the trial. To characterize treatment-mediator interaction, we calculated the controlled direct effect of the antihypertensive drug at eGFR increase and decrease by 10, 20, 30, 40, and 50%.

To quantify the extent of mediation, we calculated two metrics: proportion mediated (PM) and the proportion eliminated (PE) as follows:

Proportion mediated PM = || DE*(ME-1)/(DE*ME-1)||, where DE is a natural direct effect, and ME is a mediated effect. It captures what would happen to the effect of treatment if we disable the pathway from the treatment to the mediator, setting mediator to a single value.

Proportion eliminated PE(*m*) = (TE – CDE(*m*))/(TE-1), where TE is the total effect, and CDE(*m*) is the controlled direct effect at the level of mediator *m*. It captures what would happen to the effect of treatment on the outcome if we were to fix the mediator to the same value *M* = *m* for all persons, which is important in the case of treatment-mediator interaction.

Statistical analyses were performed using STATA MP 16.1 (StataCorp LLC, College Station, TX), and open-source code is provided at https://github.com/Tereshchenkolab/statistics.

## Results

### Study population. Relative change in eGFR during the trial

Clinical characteristics of participants stratified by the tertiles of relative eGFR change are reported in Table 1. Baseline cardiovascular risk factors and cardiovascular outcomes of ALLHAT participants in different treatment groups stratified by baseline eGFR^12, 22^, and those with different renal outcomes^14^ have been reported in detail previously.

**Table 1.** Clinical characteristics of study participants by tertiles of eGFR change

| Characteristic | Q1 eGFR change<br>(n=9,307) | Q2 eGFR change<br>(n=9,309) | Q3 eGFR change<br>(n=9,302) | P |
| --- | --- | --- | --- | --- |
| Relative eGFR change,<br>median(range) | -18.0(-94.9 to -11.01) | -0.7(-11.0 to -0.6) | +12.3(-0.55to +335) |  |
| Age(SD), y | 66.5(7.5) | 65.3(6.7) | 67.8(7.6) | <0.0001 |
| Black race, n(%) | 3,179(34.2) | 2,806(30.1) | 2,984(32.1) | <0.0001 |
| Men, n(%) | 4,815(51.4) | 5,792(62.2) | 5,221(56.1) | <0.0001 |
| HTN treated ≥ 2months, n(%) | 8,064(86.6) | 8,038(86.4) | 8,277(89.0) | <0.0001 |
| BMI(SD), kg/m<sup>2</sup> | 29.7(6.0) | 29.7(5.7) | 29.5(5.6) | 0.029 |
| Baseline SBP(SD), mmHg | 147.8(15.4) | 145.4(15.5) | 144.3(15.7) | <0.0001 |
| Baseline DBP(SD), mmHg | 84.0(10.1) | 84.3(9.9) | 83.2(10.0) | <0.0001 |
| Hx of MI/stroke, n(%) | 2,151(23.1) | 2,175(23.4) | 2,100(22.6) | 0.427 |
| Hx of revascularization, n(%) | 1,299(14.0) | 1,336(14.4) | 1,243(13.4) | 0.145 |
| Hx of ST-T or other CVD, n(%) | 2,932(31.5) | 2,878(30.9) | 3,080(33.1) | 0.004 |
| Diabetes, n(%) | 3,732(40.1) | 2,888(31.0) | 2,908(31.3) | <0.0001 |
| HDL<35mg/dL, n(%) | 995(10.7) | 1,330(14.3) | 1,249(13.4) | <0.0001 |
| Current smoking, n(%) | 2,143(23.0) | 2,054(22.1) | 1,752(18.8) | <0.0001 |
| Baseline ECG/Echo LVH, n(%) | 1,809(19.4) | 1,835(19.7) | 1,914(20.6) | 0.127 |
| Baseline total cholesterol(SD) | 216.7(43.4) | 213.9(41.2) | 215.9(41.9) | <0.0001 |
| Baseline potassium(SD) | 4.32(0.68) | 4.34(0.60) | 4.36(0.65) | 0.0007 |
| Baseline eGFR(SD) | 83.5(20.8) | 79.1(17.5) | 71.6(16.6) | <0.0001 |
| SBP change(SD), mmHg | -12.9(20.3) | -11.6(19.8) | -10.1(19.9) | <0.0001 |
| DBP change(SD), mmHg | -8.6(11.9) | -8.0(11.6) | -7.3(11.7) | <0.0001 |
| Midwest region, n(%) | 1,761(18.9) | 1,906(20.5) | 2,017(21.7) | <0.0001 |

LVH=left ventricular hypertrophy; SD=standard deviation; HTN=hypertension; BMI=body mass
index; SBP=systolic blood pressure; DBP=diastolic blood pressure; MI=myocardial infarction;
Hx=history; ST-T = major ST depression or T-wave inversion on ECG; CVD=cardiovascular
disease; CHD=coronary heart disease; HDL=high density lipoprotein

In most of the study participants, relative eGFR changes were very small (Figure 2). On average, eGFR reduced by less than one percent (median −0.7; IQR −13.3 to 8.4%). Unadjusted analysis (Table 1) showed that baseline clinical characteristics had both linear and U-shaped relationships with relative eGFR change. Alongside the relative eGFR changes (decreased – unchanged – increased eGFR), there were gradually declining baseline eGFR, baseline blood pressure, and blood pressure-lowering, but gradually rising baseline potassium. Study participants in the middle tertile of relative eGFR changes, as compared to both extremes, were slightly younger, more likely white men with a lower level of baseline total cholesterol. There were no differences in the history of MI, stroke, revascularization, or ECG/Echo-LVH between participants in different tertiles of relative eGFR change (Table 1).

**Figure 2.**
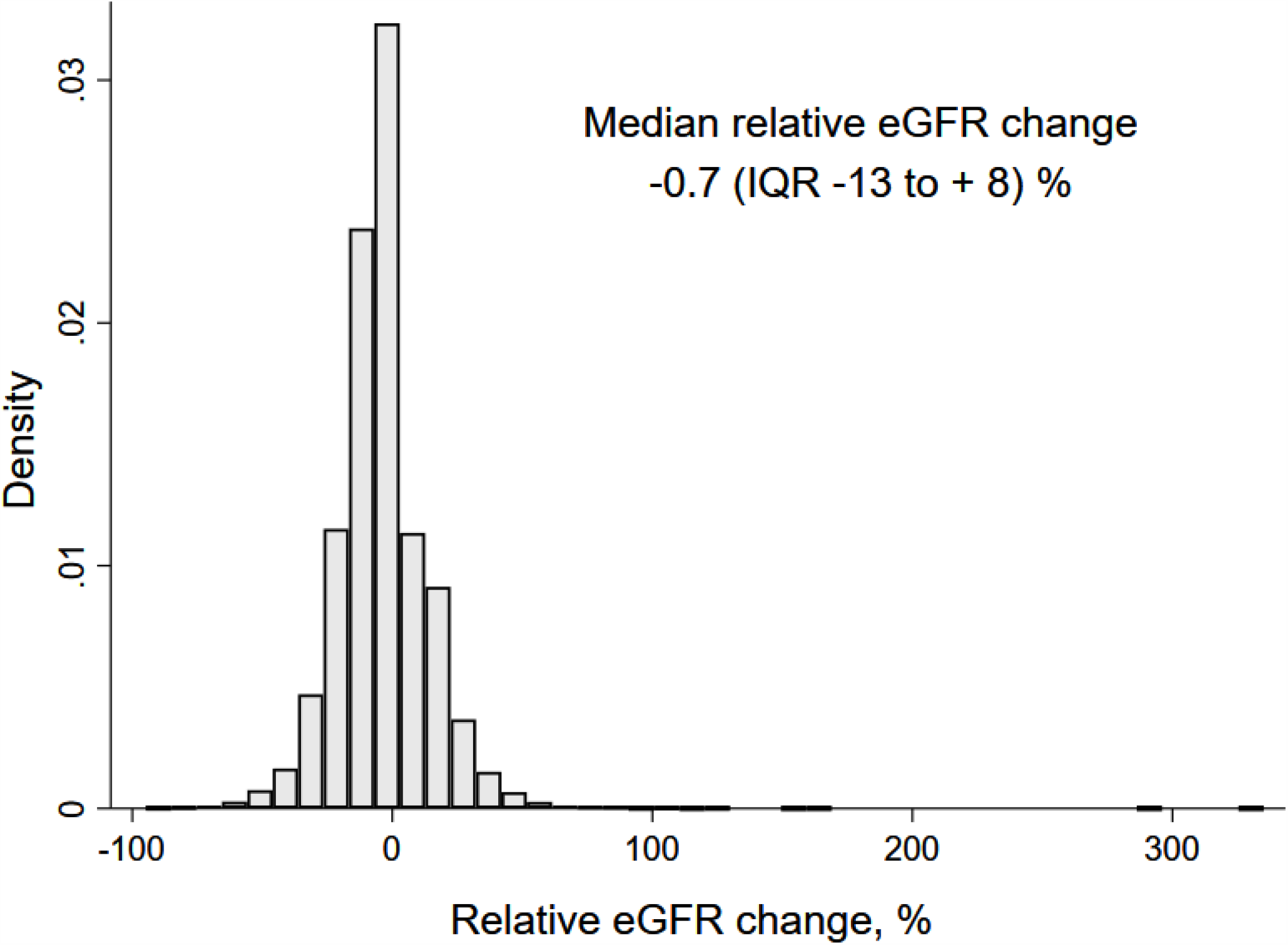
Histogram of relative eGFR change during the trial.

In a linear regression analysis adjusted for demographic characteristics (Table 2), treatment of hypertension for at least two months prior to the start of the trial, baseline HDL<35 mg/dL, Hispanic ethnicity, and Midwest or Puerto Rico residence were associated with an increase in eGFR during the trial. As expected, most of the CV risk factors (age, systolic and diastolic blood pressure, diabetes, smoking, history of MI or stroke) and Black race were associated with a decrease in eGFR during the trial (Table 2).

**Table 2.** Associations of clinical characteristics with eGFR change in linear regression

| Characteristic | Relative eGFR change(95%CI), % | P |
| --- | --- | --- |
| Age, per 1 y increase | -0.05(-0.08 to -0.02) | <0.0001 |
| Race/ethnicity: White non-Hispanic | Reference |  |
| Black non-Hispanic | -0.70(-1.18 to -0.21) | 0.005 |
| White Hispanic | +1.39(0.68-2.10) | <0.0001 |
| Black Hispanic | +1.82(0.45-3.18) | 0.009 |
| Male sex | +0.05(-0.38 to 0.48) | 0.810 |
| Hx of antihypertensive Rx ≥ 2months | +1.82(1.10-2.53) | <0.0001 |
| BMI, per 1 kg/m<sup>2</sup> increase | -0.02(-0.06 to 0.01) | 0.206 |
| Baseline SBP, per 1 mmHg increase | -0.13(-0.14 to -0.12) | <0.0001 |
| Baseline DBP, per 1 mmHg increase | -0.06(-0.08 to -0.04) | <0.0001 |
| Hx of MI/stroke | -0.87(-1.38 to -0.37) | 0.001 |
| Hx of revascularization | -1.05(-1.68 to -0.42) | 0.001 |
| Hx of ST-T on ECG or other CVD | +0.47(0.12-0.92) | 0.044 |
| Total cholesterol, per 1 mg/dL increase | +0.002(-0.004 to 0.007) | 0.540 |
| Hx of Aspirin use | -0.74(-1.18 to -0.29) | 0.001 |
| Diabetes | -3.10(-3.55 to -2.66) | <0.0001 |
| HDL<35mg/dL | +1.29(0.66-1.93) | <0.0001 |
| Current Smoking | -0.96(-1.48 to -0.43) | <0.0001 |
| Baseline potassium, per 1 mmol/L increase | +0.19 (-0.13 to 0.52) | 0.247 |
| Baseline eGFR, per 1 ml/min/1.73 m<sup>2</sup>increase | -0.22(-0.23 to -0.21) | <0.0001 |
| Baseline ECG- or echo-LVH | +0.40(-0.13 to 0.94) | 0.140 |
| Geographic region: East | reference |  |
| Midwest | +1.30(0.59-2.02) | <0.0001 |
| South | +0.48(-0.15 to 1.12) | 0.135 |
| West | +0.91(0.06 to 1.75) | 0.035 |
| Canada | +1.37(-0.15 to 2.90) | 0.077 |
| Puerto Rico | +2.34 (0.88 to 3.79) | 0.002 |

LVH=left ventricular hypertrophy; CI=confidence interval; BMI=body mass index;
SBP=systolic blood pressure; DBP=diastolic blood pressure; MI=myocardial infarction;
Hx=history; ST-T = major ST depression or T-wave inversion on ECG; CVD=cardiovascular disease; HDL=high density lipoprotein.

### Mediation of Heart Failure by eGFR changes

After a median of 3.2 years follow-up in the doxazosin group, and 5.0 years in the other three groups, there were 1,769 incident HF outcomes, including 1,359 hospitalized/fatal HF outcomes.

Overall, the relative change in eGFR during the trial mediated approximately 40% of the effect of amlodipine on the incident symptomatic HF and hospitalized/fatal HF (Table 3). For the effects of doxazosin and lisinopril, the proportion mediated hospitalized/fatal HF outcome (∼ 55 and 13%) doubled the proportion mediated symptomatic HF outcome (∼ 26 and 6%, respectively). The eGFR changes mediated more than half of the effect of doxazosin on hospitalized/fatal HF.

**Table 3.** Fully adjusted effect of antihypertensive treatment on incident heart failure (total), through eGFR changes (mediated), and independent of eGFR changes (direct).

| HF Outcome | Treatment | Total effect RR(95%CI) | Direct effect RR(95%CI) | Mediated effect RR(95%CI) | % Mediated |
| --- | --- | --- | --- | --- | --- |
| Symptomatic | Doxazosin | 1.34 (1.17-1.59) | 1.43(1.25-1.69) | 0.94(0.91 – 0.96) | 25.5 |
|  | Amlodipine | 1.44(1.27-1.67) | 1.61(1.42-1.88) | 0.90(0.86-0.93) | 38.0 |
|  | Lisinopril | 1.18(1.02-1.34) | 1.19(1.03-1.35) | 0.990(0.982-0.999) | 6.3 |
| Hospitalized / fatal | Doxazosin | 1.15(0.99-1.39) | 1.24(1.05-1.47) | 0.93(0.90-0.96) | 55.3 |
|  | Amlodipine | 1.42(1.23-1.67) | 1.60(1.38-1.86) | 0.89(0.85-0.93) | 41.9 |
|  | Lisinopril | 1.10(0.95-1.28) | 1.12(0.97-1.29) | 0.99(0.98-0.999) | 12.7 |

We observed significant treatment-mediator interaction (Figure 3). The controlled direct effect at different levels of eGFR change was considerably different. Moreover, there were significant differences in lisinopril-eGFR changes interaction for the two study outcomes (Figure 3). The decrease in eGFR during the trial was associated with a remarkable increase in relative risk of symptomatic HF for amlodipine and doxazosin treatment groups, but a slight (yet statistically nonsignificant) reduction of symptomatic HF risk for the lisinopril group. The decrease in eGFRwas associated with a notable increase in the relative risk of hospitalized/fatal HF for all treatment groups. The increase in eGFR during the trial was associated with a slight reduction of the relative risk of hospitalized/fatal HF for all treatment groups, and reduction of symptomatic HF risk for doxazosin and amlodipine groups, but trivial increase in symptomatic HF risk for lisinopril group.

**Figure 3.**
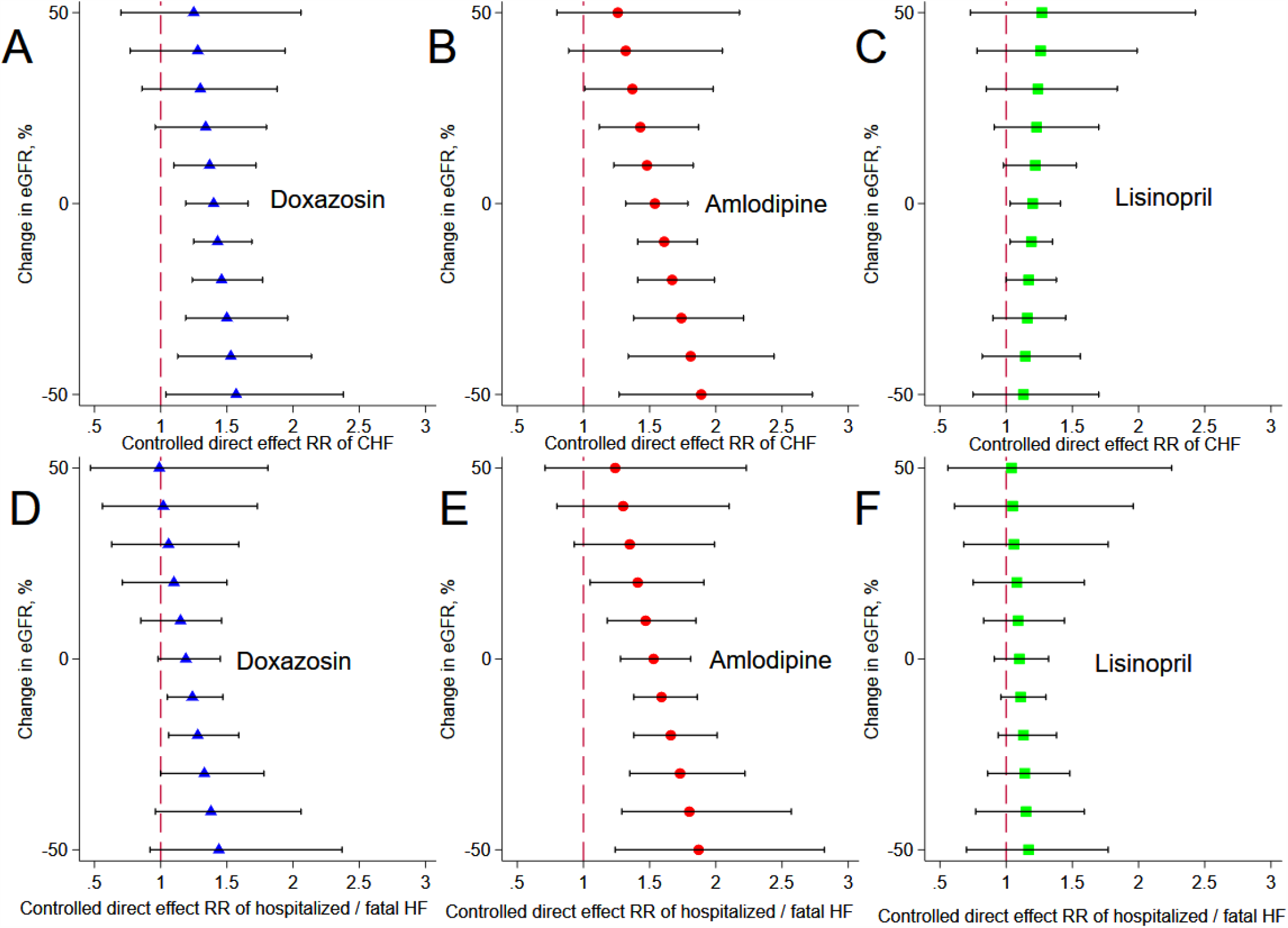
Controlled direct effect of doxazosin (**A**,**D**), amlodipine (**B**,**E**), and lisinopril (**C**,**F**) at eGFR increase and decrease by 10, 20, 30, 40, and 50%.Relative Risk of incident symptomatic congestive HF (**A, B, C**) and hospitalized / fatal HF (**D, E, F**). All models were adjusted for age, sex, race/ethnicity, baseline systolic and diastolic blood pressure, in-trial systolic and diastolic blood pressure lowering, length of antihypertensive treatment before enrollment, ECG/Echo-LVH, history of MI, stroke, or other CVD, coronary revascularization, major ST-T changes, HDL<35 mg/dL, BMI, smoking, diabetes, use of aspirin, participation in the lipid-lowering ALLHAT trial, baseline total cholesterol, eGFR, and potassium, and geographic region.

Proportion eliminated (Figure 4) was remarkably high for all treatment groups and both outcomes. For patients in doxazosin arm, the effect of eGFR changes on hospitalized/fatal HF was twice larger than on symptomatic HF. Reduction in eGFR by at least 30% explained 50% or more of symptomatic HF (Figure 4A) and was the main and only cause of hospitalized/fatal HF (Figure 4D). “Equipoise” was reached for patients treated with doxazosin who achieved slight improvement in eGFR, by 10%, - it was inconsequential in terms of HF development. An increase in eGFR by 50% explained 28.6% of the symptomatic HF risk reduction and 100% of hospitalized/fatal HF.

**Figure 4.**
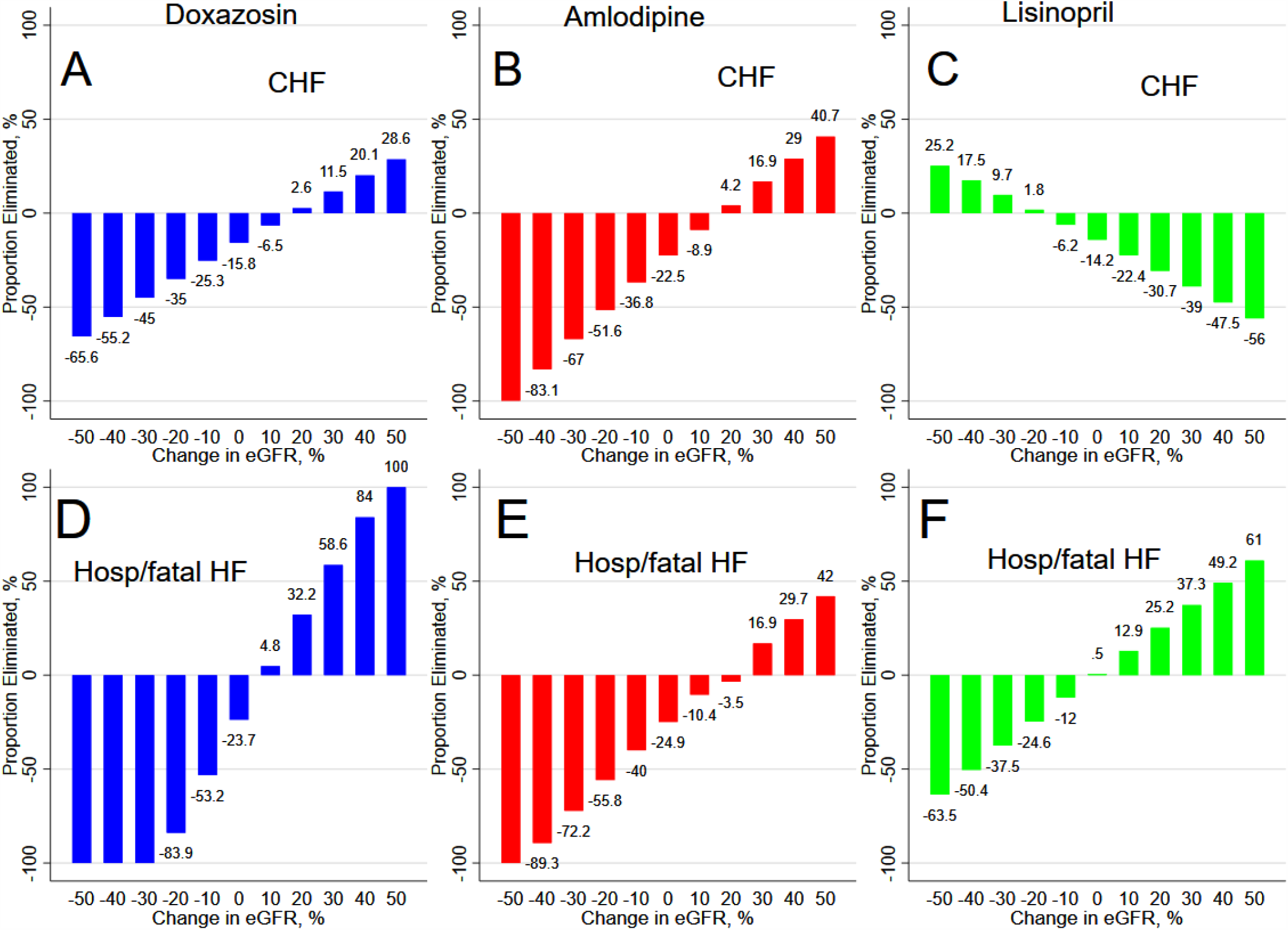
Proportion eliminated of the effect of doxazosin (**A, D**), amlodipine (**B, E**), and lisinopril (**C, F**) on incident symptomatic congestive HF (**A, B, C**) and hospitalized / fatal HF (**D, E, F**) at eGFR increase and decrease by 10, 20, 30, 40, and 50%. All models were adjusted as described in Figure 3 legend.

For patients in the amlodipine arm, the effect of eGFR changed on both outcomes was similar. Reduction in eGFR by at least 20% explained more than 50% of both symptomatic and hospitalized/fatal HF (Figure 4B and E). Even if eGFR did not change, unchanging eGFR was responsible for approximately 25% of HF in patients treated with amlodipine. An equipoise required a 20% increase in eGFR – in such cases, eGFR change did not impact HF development. An increase in eGFR by 50% explained approximately 40% risk reduction of both symptomatic and hospitalized/fatal HF mechanisms.

For patients in lisinopril arm, eGFR changes had an opposite effect on symptomatic versus hospitalized/fatal HF outcomes. Reduction in eGFR by at least 40% explained more than 50% of increased risk in hospitalized/fatal HF (Figure 4F) but also explained an 18-25% *reduction* of symptomatic HF risk (Figure 4C). Unchanged eGFR did not contribute to hospitalized/fatal HF development. However, unchanged eGFR explained 14.2% of the increase of symptomatic HF risk. An equipoise - no effect by eGFR changes on symptomatic HF outcome-required eGFR reduction by 20%. An increase in eGFR by 50% explained 61% of the hospitalized/fatal HF risk reduction, and 56% of the symptomatic HF risk *increase*.

## Discussion

This causal mediation analysis of the largest randomized controlled trial of antihypertensive treatment revealed that in hypertension patients with normal baseline kidney function, eGFR change was the main mechanism of incident HF. Even as small as 10% relative eGFR changes explained up to 50% of HF mechanisms. Large (±50%) eGFR change was frequently the main and only HF mechanism. There were differences in the degree of treatment-mediator interaction - between treatment groups and between two HF outcomes. In amlodipine arm, the effects of eGFR change on both symptomatic and hospitalized/fatal HF were similar, whereas, in doxazosin arm, the effect of eGFR change on hospitalized/fatal HF was approximately twice larger than on symptomatic HF. Remarkably, eGFR change had an opposite effect on symptomatic versus hospitalized/fatal HF in lisinopril arm. A decline in eGFR mediated increase in hospitalized/fatal HF, but reduction in symptomatic HF relative risk. Respectively, relative eGFR increase mediated increased risk of symptomatic HF, but the lessening of hospitalized/fatal HF risk. Appropriate use of antihypertensive medications causing beneficial eGFR changes can help to eliminate a substantial proportion of HF risk.

### Is heart failure a kidney disorder?

Observational cohort studies provided the foundation for the cardiorenal syndrome definition with its characteristic feature of tangled interconnected concomitant presence of kidney dysfunction, and HF.^23^ Hypertension is the leading risk factor of both CKD and HF.^24^ Our causal mediation analysis of the largest randomized controlled trial supported the notion of HF as a kidney disorder.^25^ While hypertension is one of the major risk factors for HF^26^, blood pressure-lowering mediated only up to 13% of the effect of the antihypertensive medications on incident HF.^18^ We found that the kidney function mechanism explains a large portion of the effect of antihypertensive medications on HF. Thus, further refinement of interventions targeting this mechanism can be suggested to increase the magnitude of the effect on HF prevention.

Hypertension is characterized by arterial stiffening and endothelial dysfunction, which can lead to nephrons loss, compensatory hyperfiltration of the remaining nephrons in an attempt to maintain GFR, and eventually, renal dysfunction.^27, 28^ Concomitant volume retention can lead to the volume-overloaded state, increased central venous pressure, and eventually, pulmonary hypertension and congestion, and symptomatic HF.^29^ Endothelial dysfunction as a result of renal impairment facilitates myocyte stiffening and subsequent fibrosis, and provoke the inflammatory cascade and oxidative stress, predisposing to myocardial dysfunction.^30, 31^ Our finding suggests that HF prevention cannot be successful without targeting the main HF mechanism – kidney dysfunction, which is supported by stalled progress in HF prevention for several decades.^32^

An average ALLHAT participant had a normal or mildly reduced baseline kidney function and a low risk for the development of ESKD.^12, 33^ Only approximately 20% of ALLHAT participants had CKD (eGFR < 60 mL/min/1.73m^2^) at the time of randomization. Of note, ALLHAT excluded individuals with a history of symptomatic or hospitalized HF and LVEF < 35%. The development of ESKD, which was the prespecified secondary outcome was not different between treatment groups.^10, 14^ Thus, the ALLHAT study population was well-suited for the study of kidney dysfunction as a hypothesized HF mechanism. Importantly, we assessed dynamic changes in eGFR during the trial and applied rigorous causal mediation analysis adjusted for confounders. Altogether the rigorous study design, study population, and analytical approach support the main study conclusion about kidney dysfunction as the major mechanism of HF development in hypertensive patients.

There is an ongoing debate about whether or not eGFR reduction always reflects kidney dysfunction. In acute HF, in aggressively diuresed HF patients, transient creatinine increase may represent potentially benign hemodynamically driven eGFR reduction. However, we assessed the eGFR difference over at least two years time period. There is also a belief that while the use of ACEIs may lead to eGFR reduction in short-term, long-term ACEIs exert additional CV benefit via the preservation of kidney function. Results of our study question this assumption. Indeed, consistently with the common belief, a slight reduction in eGFR in lisinopril arm was associated with a relative reduction in symptomatic HF risk. Nevertheless, at the same time, it associated with an increased risk of hospitalized/fatal HF. Mediation of hospitalized/fatal HF by eGFR changes had the same direction in all treatment groups: the greater eGFR reduction, the higher risk of hospitalized/fatal HF. An “unconscious bias” embedded in a clinical judgment could possibly explain our findings. Striving to differentiate symptomatic HF from volume overload due to kidney dysfunction, physicians (ALLHAT investigators) were more likely to attribute a patient’s symptoms to HF and diagnose symptomatic HF if a patient was presented with improving eGFR. And in the opposite, if a patient was presented with eGFR worsening, physicians were more likely to attribute the same symptoms to volume overload due to kidney dysfunction and less likely to diagnose symptomatic HF. The results of this study suggest that attempts to differentiate HF from kidney dysfunction can be misleading and put emphasis on the improvement of a “heart pump function” while the focus should be on the improvement of kidney function.

### Strengths and Limitations

ALLHAT is the largest RCT of antihypertensive treatment, allowing unbiased mediation analysis, strengthening two major assumptions of mediation analysis. The development of ESKD was a prespecified secondary outcome in ALLHAT^9^, which ensured the careful collection of dynamic eGFR data. Randomization eliminated exposure-outcome and exposure-mediator confounding.

However, limitations of this study should be taken into account. While we adjusted for known common causes of eGFR changes and incident HF, unmeasured confounding can affect this study’s estimates. ALLHAT enrolled high-risk HTN patients, and the results of this study may not be generalizable to a lower-risk population. ALLHAT also did not report urine albumin in the patient population, which hindered us from using the degree of albuminuria, an important marker for CKD in this study.

## Data Availability

We used the ALLHAT dataset that is publicly available from the National Heart, Lung, and Blood Institute, via BioLINCC.

## Acknowledgment

This manuscript was prepared using ALLHAT Research Materials obtained from the NHLBI Biologic Specimen and Data Repository Information Coordinating Center.

## Funding Sources

This work was partially supported by HL118277 (Tereshchenko). The ALLHAT study was supported by the National Heart, Lung, and Blood Institute (NO1-HC-35130). ALLHAT investigators received contributions of study medications supplied by Pfizer (amlodipine and doxazosin), AstraZeneca (atenolol and lisinopril), Bristol-Myers Squibb (pravastatin), and financial support provided by Pfizer.

## Disclosures

None

## Notes

### Competing Interest Statement

The authors have declared no competing interest.

### Clinical Trial

NCT00000542

### Author Declarations

The Oregon Health & Science University Institutional Review Board reviewed the study and determined the deidentified nature of the publicly available dataset.

